# An integrated compositional approach to investigating the full effect of dietary intake on health

**DOI:** 10.1101/2022.10.20.22281295

**Authors:** Maria Léa Corrêa Leite

## Abstract

Nutritional epidemiology studies aim to investigate the impact of diet on disease risk, and their findings inform subsequent dietary recommendations. The issue of whether foods or nutrients should be the unit of exposure in such studies is a topic of debate. However, although nutrient-based analyses can elucidate biological mechanisms, they are technically problematic; furthermore, although foods have been considered the most suitable unit of dietary exposure because “we eat food, not nutrients” and the findings of food-based analyses can be easily translated into dietary advice, there is a greater risk of confounding and/or reverse causation because food selection and consumption are closely related to socio-economic and behavioural factors. This note describes an integrated approach that combines the positive aspects of nutrient-and food-based analyses. Individual dietary exposure is characterised by its overall macro-and micronutrient compositions and, in line with the compositional nature of dietary data, methods based on log-ratio transformations of nutrient compositions are used in diet/outcome association analyses that provide fully adjusted isocaloric estimates of the impact of specific nutrient balances on health outcomes. These estimates can then be combined with the nutrient compositions of selected foods to produce outcome-related scores which, as they are defined regardless of a subject’s food selection/consumption, are not subject to confounding or reverse causation, and therefore have important implications for public health professionals. Data coming from an Italian population-based study were used to illustrate the approach in the context of diet/serum uric acid relationships, and some resulting new insights were briefly discussed.

## 1. Introduction

The overall aims of nutritional epidemiology research include investigating the relationships between diet and disease risk that may have an impact on public health as they inform the development of specific dietary recommendations among measures designed to prevent certain diseases.

The last two decades have seen substantial debate as to whether foods or nutrients should be considered the fundamental unit of exposure in such studies: it is acknowledged that nutrient-based analyses are suitable means of elucidating the biological mechanisms underlying the associations between diet and disease risk, but has been claimed that foods are more appropriate because nutrients are not consumed in isolation but in the context of complex food matrices.

Moreover, from a practical point of view, nutrient-based analyses raise technical problems such as collinearities that make it difficult to obtain reliable estimates of the effect of a single nutrient, whereas the results of food-based analyses can be directly translated into dietary advice.

However, even when adopting a food-based approach, researchers wonder about biological mechanisms and inevitably try to interpret the results for a given food in terms of its nutrient components: for example, by attributing the observed beneficial effects of diets rich in fruit and vegetables on the risk of cardiovascular disease risk to the presence of certain vitamins and antioxidants, or those of olive oil to its high levels of monounsaturated fatty acids. However, it is somewhat disconcerting to note that beneficial effects of these constituents have often not been confirmed in randomised trials.

The simple statement used to justify the choice of food as the unit of analysis (“we eat food, not nutrients”) is in itself suggestive of the frequently unrecognised drawback when performing food-based analyses: i.e., the greater chance of confounding. Food selection and consumption are closely related to socio-economic and behavioural factors that can directly or indirectly influence disease risk, and adjustments for measures of these potential confounders by means of standard statistical tools are probably inadequate or insufficient to fully capture the extent of the complex ways in which social behaviours confound the associations between foods and disease. Moreover, food choices may be driven by a subject’s self-perceived health status, thus making the occurrence of reverse causation more likely.

This note is based on the assumptions that diets consist of the set of foods consumed and that our biological systems do not face one single nutrient or food matrix at a time, but the whole-nutrient composition of a combination of various food matrices. It therefore seems to be more appropriate to characterise individual dietary exposure on the basis of its overall macro-and micronutrient compositions.

Any composition is represented by parts and their relationships, and conveys substantially relative information that can be properly represented in the form of ratios [1]. We have previously described the application of ratio-based compositional data analysis to nutritional studies [2]. This approach does not consider isolated and unrealistic variations in single nutrients, but provides association estimates based on the *balances* of specific nutrients while suitably accounting for total energy intake and the proportional relationships between all of the other dietary components.

Food is undoubtedly the preferred unit of communication because there is a clear practical advantage in offering dietary guidance based on food selection, but the question is how to apply nutrient balance-related inferences at the level of foods in order to inform dietary recommendations. A food is per se a nutrient composition: the concept of a food matrix is, in effect, fully comparable to the definition of a composition. We have previously proposed a means of defining health-related food scores that combine association estimates relating to nutrient balances with the nutrient compositions of the foods themselves [3]. Such scores are not subject to the counfounding described above because they are not based on food selection/consumption.

This note describes an integrated approach to evaluating the relationships between dietary intakes and health outcomes that combines the above mentioned procedures.

## 2. Methods

### 2.1 Log-ratio transformations

Different expressions of log-ratios have been proposed for the analysis of compositional data. Log-ratio transformations produce new variables that can be included as covariates in standard regression models. Given a D-part composition (x_1_, x_2_, …,x_D_), the isometric log-ratio (ilr) transformation [4] in its particular form of *balances*, consists of orthogonally decomposing the parts of a whole into non-overlapping subgroups and representing their relationships. One way of constructing orthonormal balances is to use sequential binary partition (SBP) as a bifurcating tree: the parts of a composition are successively and hierachically split into two groups until all of the groups have a single part. At each of the D-1 steps required to complete the partition, a generic balance is defined as the orhonormal log-ratio of the geometric mean of each group of parts:

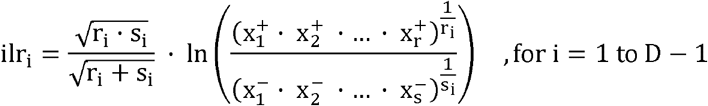

where the square root coefficient rapresents the nomalising constant, and r and s are respectively the number of parts in the numerator (x^+^) and the number of parts in the denominator (x^-^).

One particular choice of partition defined as:

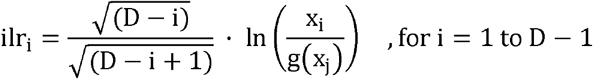

has been proposed [5] where g (x_j_) is the geometric mean of the compositional parts for j=i+1 to D.

The particular characteristic of this partition is that the first new variable (ilr_1_) captures all of the relevant information about the compositional part x_1_, and is thus called the *pivot balance* [6]. This special balance expresses the *relative dominance* of the part in the numerator over the other compositional parts. A number of D sets of D-1 ilr(s) are defined, each of which has a different compositional part in the numerator of the pivot balance.

The removal of the normalising constant, and consequently the reduction of the orthonormality of the log-contrasts to simple orthogonality, has been proposed [7] in order to enhance the interpretability of the results of regression analyses involving ilr-balances as explanatory variables.

If researcher is interested in evaluating the impact of the relative dominance of each compositional part, it may be convenient using the additive log-ratio transformation which involves the division of each D-1 component by one that is arbitrarily chosen, for example the last:

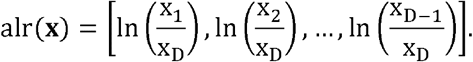

Actually, it has been shown [8] that alr and simplified (orthogonal rather than orthonormal) pivot balances are explanatory equivalents. Using alr transformation as predictors and only two runs leads to the same result as the pivot approach which requires a number of runs that is equal to the number of components in the composition. We have previously described the use of different log-ratio transformations of dietary compositions and discussed in detail the interpretative implications of using them as explanatory variables [2].

Suppose we have defined a number (I) of dietary balances relating to macro-and micronutrient compositions, and fitted a regression model that has a given health outcome (HO) as a response variable whose expected value is:

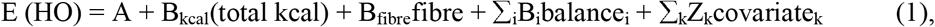

where the B coefficients (B_kcal_, B_fibre_ and B_i_, for i=1 to I) express the association between the HO and the dietary variables, and Z_k_ relates to the k control covariates.

### 2.2. Outcome-related scores for foods

The following step of the analysis consists of a procedure described in detail elsewhere [3] that involves defining scores for some selected foods on the basis of the weighted average of the diet-HO association estimates represented by B-coefficients such as those in (1). The score for a generic food f (Sc_f_) for i=1 to (I+2) is:

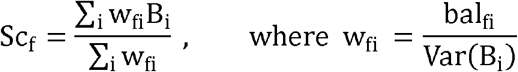

The B-coefficients are obtained from equations that are similar to (1) but containing special *balances*, as explained below. These special balances are then also calculated for each food (bal_fi_) on the basis of their nutrient contents. The weights (w_fi_) are then directly related to the nutrient balances of the foods, and inversely related to the variances of the regression coefficients (Var(B_i_)), thus privileging more precise estimates.

Unlike the log-ratio definitions relating to the overall nutrient composition of a diet, it may be a problem when the calculation involves nutrient compositions of a given food because of the presence of zero observations for which logarithms are not suitable. Rounded zeroes (i.e. values below the limit of detection) may be simply replaced by an appropriately small value, whereas essential zeroes, which indicate the real absence of a part (nutrient) from a composition (food matrix), can be treated by exploiting the flexibility of the sequential binary partition procedure. The procedure has been described in detail elsewhere [3]. Briefly, we begin by dividing the foods into subgroups on the basis of similarities in their nutrient composition, and then identify the partition that makes it possible to “isolate” the lacking nutrient and define balances without losing any other information.

In order to illustrate the key idea underlying the procedure, consider a simple case of a 5-nutrient composition (n_1_, n_2_, …,n_5_) and two alternative partitions (A and B) represented by the following sign matrices in which the plus sign (+) indicates that that part of the balance is assigned to the numerator, the minus sign (-) indicates that it is assigned to the denominator, and zero (0) indicates that it is not involved in that particular balance:

| Step | $n_1$ | $n_2$ | $n_3$ | $n_4$ | $n_5$ | Orthogonal balances |
| --- | --- | --- | --- | --- | --- | --- |
| 1 | + | + | - | - | - | $\ln((n_1 * n_2)^{(1/2)} / (n_3 * n_4 * n_5)^{(1/3)})$ |
| 2 | + | - | 0 | 0 | 0 | $\ln(n_1/n_2)$ |
| 3 | 0 | 0 | + | - | - | $\ln(n_3/(n_4 * n_5)^{(1/2)})$ |
| 4 | 0 | 0 | 0 | + | - | $\ln(n_4/n_5)$ |

B
| Step | $n_1$ | $n_2$ | $n_3$ | $n_4$ | $n_5$ | Orthogonal balances |
| --- | --- | --- | --- | --- | --- | --- |
| 1 | - | - | - | + | - | $\ln(n_4/(n_1 * n_2 * n_3 * n_5)^{(1/4)})$ |
| 2 | + | + | - | 0 | - | $\ln((n_1 * n_2)^{(1/2)} / (n_3 * n_5)^{(1/2)})$ |
| 3 | + | - | 0 | 0 | 0 | $\ln(n_1/n_2)$ |
| 4 | 0 | 0 | + | 0 | - | $\ln(n_3/n_5)$ |

Let us then suppose the case of a particular food which is devoid of the nutrient n_4_. By using the partition A, the balance corresponding to the step 2 is the only calculable because it does not involve n_4_. Alternatively, the partition B allows to capture (and isolate) all of the information relative to the lacking nutrient into the first balance which can be ignored (or put equal zero) when calculating the score for this food, without losing any information regarding the other components of the nutrient composition. Note that the nutrient n_4_ does not appear in the successive balances, and therefore the partition B is the appropriate choice.

It is worth noting that the different partitions represent orthogonal rotations of the system of balances, and that all of the regression equations contains the same number of explanatory variables. This means that all of the other terms and performance measures of the model remain exacly the same in the different runs, thus assuring the comparability of the food scores regardless of the model from which the B-coefficients are taken.

## 3. Example: serum uric acid (SUA) levels as the health outcome

The example makes use of data from the Italian Bollate Eye Study [9], a population-based study of subjects aged 40-74 years conducted in 1992 and 1993. The dietary habits of the participants were assessed by means of a food-frequency questionnaire, and their mean daily nutrient intakes were calculated using the food compositional database compiled for epidemiological studies in Italy [10].

The example below considers the serum uric acid (SUA) levels as the health outcome. Hyperuricemia is the precursor of gout and may be a strong risk factor for cardiovascular and renal diseases, metabolic syndrome and diabetes. On the other hand, low levels of SUA have been associated with a higher prevalence of some neurological diseases. The relationship between diet and SUA levels, although widely investigated, is still unclear. However, broadly hyperuricemia has found positively associated with intakes of meats, seafoods and alcohol and negatively with dairy products and somewhat with coffee consumption.

Linear regression models were designed with the levels of SUA as the dependent variable and, as the concentration units (mg/mL) indicate compositional information, their values were logged in order to allow an estimate of relative changes. This analysis considered three dietary compositions: a nine-part macronutrient (energy source) composition; a six-part mineral composition; and a nine-part vitamin composition. In addition to the independent dietary variables of ln(kcal), ln(fibre), and I macro-and micronutrient balances, the models also included terms for the K covariates of age, gender, smoking, practising sport, television watching time, and diuretic use:

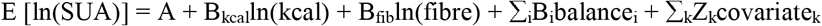

### 3.1. Results and some comments

The study population consisted of 1,162 subjects (593 men and 569 women), and the mean values (standard deviation) of SUA levels (in mg/dl) were 5.30 (1.26) among the men and 4.12 (1.12) among the womem. The tables show the results of the regression analyses. The coefficients represent the expected change in the dependent variable when the related balance is multiplied by an e-factor of 2.7, and as the balances were mutually adjusted, the nutrients appearing in the numerator of the balances increased by a common factor, and those in the denominator decreased by a common factor.

Table 1 shows the results for the macronutrients from two alternative approaches: model 1 which included the orthogonal balances defined following the natural clustering of the macronutrients, and model 2 which included the alr(s) that made it possible to evaluate the impact of individual relative dominances. Consistently with other studies [11,12], the relative dominance of alcohol over other sources of calories is positively and significantly associated with SUA levels.

**Table 1.** Linear regression coefficients (B) and standard errors (SE) of serum uric acid levels [ln(mg/ml)] in relation to total energy and fibre intake and the balances of the source of the calories (alcohol and macronutrients).

| Model 1 |  |  | Model 2 |  |  |
| --- | --- | --- | --- | --- | --- |
|  | B (SE) | P-value |  | B (SE) | P-value |
| Total energy [ln(kcal)] | 0.066 (0.024) | 0.006 | Total energy [ln(kcal)] | 0.066 (0.024) | 0.006 |
| Total fibre [ln(mg)] | 0.076 (0.056) | 0.178 | Total fibre [ln(mg)] | 0.076 (0.056) | 0.178 |
| *Orthogonal balances:* |  |  | *Relative dominances:* |  |  |
| Alcohol vs macronutrients | 0.034 (0.009) | 0.000 | Alcohol vs macronutrients (mac.) | 0.034 (0.009) | 0.000 |
| Fats vs proteins and carbohydrates | 0.052 (0.047) | 0.267 | Animal proteins vs other mac. | -0.019 (0.045) | 0.668 |
| Proteins vs carbohydrates | 0.030 (0.069) | 0.663 | Vegetable proteins vs other mac. | 0.024 (0.078) | 0.761 |
| Animal vs vegetable proteins | -0.022 (0.048) | 0.656 | Starches vs other mac. | -0.065 (0.054) | 0.228 |
| Starches vs simple sugars | -0.037 (0.030) | 0.226 | Simple sugars vs other mac. | 0.009 (0.029) | 0.762 |
| Saturated vs unsaturated fats | -0.014 (0.040) | 0.732 | Saturated fats vs other mac. | -0.001 (0.042) | 0.985 |
| Monounsaturated vs polyunsaturated fats | -0.061 (0.030) | 0.047 | Monounsaturated fats vs other mac. | -0.043 (0.038) | 0.256 |
| n-6 linoleic vs n-3 alpha-linolenic acids | -0.022 (0.034) | 0.525 | n-6 Linoleic acid vs other mac. | 0.026 (0.041) | 0.528 |
|  |  |  | n-3 Alpha-linolenic acid vs other mac. | 0.070 (0.040) | 0.081 |

The models also included terms for gender, age, practising sport, television watching time, smoking, diuretic use, and micronutrient (mineral and vitamin) balances.

Table 2 shows the results for the dietary vitamins and minerals obtained from the model including the alr(s), and so are expressed in terms of their individual relative dominances, among which the inverse association of SUA levels with the relative dominance of vitamin B6 stood out from the other estimates. The 2.7 factor increase in the vitamin B6 simplified pivot balance (shown in such a way that this vitamin increased and all of the others decreased by a common factor) was related to the expected multiplicative change in SUA levels of *e*(−0.161) = 0.851, an approximately 15% reduction. The few nutritional studies that have investigated the association between vitamin B6 (pyridoxine) and SUA levels have not reported any association between vitamin B6 intake and the risk of hyperuricemia [13] or between the serum levels of vitamin B6 and those of UA [14]. However, the result shown in the example is consistent with experimental evidence showing that vitamin B6 greatly inhibits the activity of xanthine oxidase, the enzyme responsible for the last steps of the purine catabolism that promotes the formation of uric acid [15,16].

**Table 2.** Linear regression coefficients (B) and standard errors (SE) of serum uric acid levels [ln(mg/ml)] in relation to the individual relative dominances defined for the dietary vitamin and mineral compositions.

| Vitamins |  |  | Minerals |  |  |
| --- | --- | --- | --- | --- | --- |
| *Relative dominances* | B (SE) | P-value | *Relative dominances* | B (SE) | P-value |
| Thiamin *vs* other vitamins | 0.046 (0.065) | 0.482 | Sodium *vs* other minerals | 0.024 (0.019) | 0.208 |
| Riboflavin *vs* other vitamins | 0.049 (0.065) | 0.449 | Potassium *vs* other minerals | 0.004 (0.081) | 0.963 |
| Niacin *vs* other vitamins | 0.047 (0.061) | 0.434 | Calcium *vs* other minerals | -0.020 (0.042) | 0.633 |
| Vitamin B6 *vs* other vitamins | -0.161 (0.078) | 0.040 | Phosphorus *vs* other minerals | -0.031 (0.097) | 0.747 |
| Folates *vs* other vitamins | 0.005 (0.057) | 0.925 | Iron *vs* other minerals | -0.008 (0.063) | 0.896 |
| Vitamin D *vs* other vitamins | -0.001 (0.016) | 0.962 | Zinc *vs* other minerals | 0.032 (0.079) | 0.684 |
| Vitamin E *vs* other vitamins | -0.020 (0.050) | 0.687 |  |  |  |
| Vitamin A *vs* other vitamins | 0.048 (0.028) | 0.079 |  |  |  |
| Vitamin C *vs* other vitamins | -0.014 (0.029) | 0.625 |  |  |  |

The models also included terms for gender, age, practising sport, television watching time, smoking, diuretics use, total energy and fibre intake and the source of the calories (alcohol and macronutrients) balances.

Figure 1 shows the uricogenic scores of the selected foods, listed from the highest (at the top) to the lowest score. Note that the scores calculated for spirits > wine > beer reflect their alcohol content, but other studies have found that beer leads to a greater increase in SUA levels than that induced by other alcoholic drinks, and that the consumption of red wine is not associated with SUA levels [11,17]. As SUA is the end product of purine degradation, it has been suggested that the higher purine content of beer may explain its greater effect on SUA levels, but this has been challenged by the findings of studies showing that neither the consumption of purine-rich vegetables nor total protein intake (it is worth remembering that protein-rich diets tend to contain large quantities of purines) is associated with an increased risk of hyperuricemia and gout [18-20]. However, given that some studies have consistently shown that wine drinkers have healthier diets and behaviours than other drinkers [21-23], it seems more reasonable to conjecture that confounding may have masked the uricogenic effect of wine.

**Figure 1.**
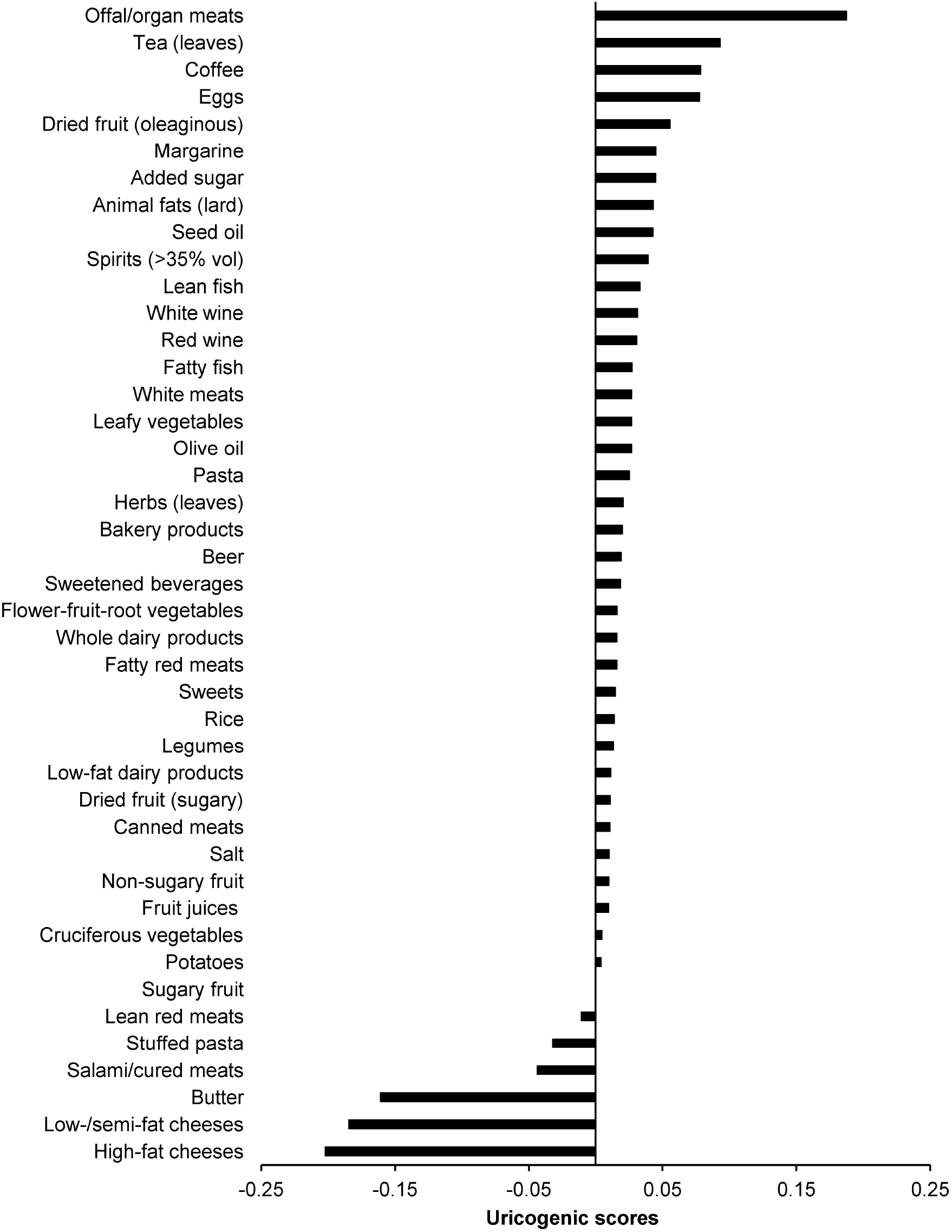

Tea and coffee respectively had the second and third highest uricogenic scores, which conflicts with the findings of some studies that have shown that coffee lowers SUA levels [24], whereas tea does not seem to have any effect [25]. As observational studies have reported inverse relationships between coffee consumption and metabolic disturbances closely related to hyperuricemia, such as insulin resistance, it has been suggested that increased insulin sensitivity may be the explanation [26]. However, this hypothesis has been contrasted by Mendelian Randomisation studies showing that coffee consumption increases the risk of diabetes and obesity [27], or is not associated with either condition [28]. It has therefore been suggested that the most likely causes of the observational findings are unmeasured confounding factors or reverse causation [28].

## 4. Conclusions

This note describes an integrated approach to investigating the relationships between health outcomes and diet that takes into account all of the available dietary infomation and makes it possible to make inferences concerning the health effects of variations in specific nutrient balances. Furthermore, on the basis of these inferences and the nutrient contents of selected foods, it is possible to define scores that allow foods to be ranked on the basis of their potential impact on health outcomes and, as the scores are calculated regardless of the habitual individual consumption of the foods themselves, they are not subject to the greater risk of confounding characterising food-based analyses. This approach satisfies two important objectives of nutritional epidemiology studies: it provides data that can improve our understanding of biological mechanisms and inform the definition of dietary recommendations in terms of food choices. For example, ranking foods on the basis of their uricogenic potential can facilitate the tailoring of such recommendations to the clinical characteristics of individuals insofar as the association of SUA with health risk is biphasic: i.e. high SUA levels are associated with cardiovascular and metabolic diseases, whereas low SUA levels are associated with neurodegenerative diseases.

One limitation of the described analysis is that it was based on non-exhaustive nutrient compositions, which means the procedure can be improved by increasing the number of dietary constituents. Given a suitable sample size, the more complete the nutrient compositions, the more accurate the calculation of the food scores, and the more finely adjusted the balance-related associations.

In summary, this note describes an integrated approach to investigating the effects of diet on health that has the advantage of being based on the compositional nature of dietary data. Isocaloric association estimates concerning nutrient balances can be extensively and finely adjusted by means of models that include terms for log-ratios expressing the proportional relationships among all of the components of a diet. Furthermore, the procedure allows the definition of food scores that are unrelated to a subject’s dietary habits and based on the intrisic proprieties of the food themselves and, as the scores indicate the potential influence of foods on health outcomes, they may have important implications for professionals making public health recommendations and giving dietary advice.

## Data Availability

All data produced in the present study are available upon reasonable request to the authors

## Funding

None.

## Conflict of interest

None.

## Figure legend

Weighted averages of the regression coefficients representing estimates of the association between dietary nutrient balances and serum uric acid levels. The weights are given by the nutrient balances within each food, and are inversely related to the variance of the regression coefficient estimates.

## Notes

### Competing Interest Statement

The authors have declared no competing interest.

### Funding Statement

This study did not receive any funding

### Author Declarations

Ethics Committee of the National Research Council (CNR) gave ethical approval for this work.

